# Prevalence and determinants of fertility care-seeking among women with delayed conception in Northern India: a cross-sectional study

**DOI:** 10.1101/2025.11.12.25340115

**Authors:** Barsha Gadapani Pathak, Gitau Mburu, Ndema Habib, Rita Kabra, James Kiarie, Ranadip Chowdhury, Neeta Dhabhai, Sarmila Mazumder

## Abstract

Infertility affects a substantial proportion of couples in low– and middle-income countries, yet access to fertility care remains limited. In India, research has primarily focused on psychosocial and biomedical aspects of infertility, with limited attention to care-seeking behaviours. Understanding the determinants of care-seeking is essential to promote equitable access to infertility services. We conducted a cross-sectional study among 1,530 married women, aged 18–30 years, intending to conceive and living in low-to middle-socioeconomic urban neighbourhoods of Delhi who had not conceived after 18 months of participation in a preconception randomized-control trial. Structured interviews captured sociodemographic characteristics, fertility intentions, psychosocial experiences, and care-seeking actions. Care-seeking was categorized as formal (health system providers), informal (family or traditional healers), or mixed. Overall, 69.9% of women sought some form of help for delayed conception and, among them 85.5% consulted both formal and informal sources. Multivariable logistic regressions identified predictors of care-seeking where longer duration of trying to conceive (adjusted odds ratio [aOR] 1.6 per year, 95% CI 1.4–1.7), perceiving conception as taking too long (aOR 3.1, 95% CI 2.1–4.5), feeling isolated (aOR 1.7, 95% CI 1.2–2.4), emotional abuse from husbands (aOR 1.7, 95% CI 1.2–2.4) or family members (aOR 1.6, 95% CI 1.2–2.2), and heavy menstrual bleeding (aOR 1.5, 95% CI 1.0–2.2) were associated with greater odds of care-seeking, while regular menstrual cycles were associated with lower odds (aOR 0.6, 95% CI 0.4–0.9). The negative association of regular cycles with care-seeking was significantly modified by social isolation (interaction aOR 3.78, 95% CI 1.20–11.89). The most common reason for not seeking care was the perception of not having a problem. Nearly seven in ten women with delayed conception sought some form of help, often through mixed care pathways. Findings highlight the need to integrate psychosocial support and fertility education into accessible, affordable infertility services within the public health system.

## Introduction

Infertility is a disease of the male and female reproductive system defined as the failure to achieve a clinical pregnancy after 12 months of regular unprotected sexual intercourse (1, 2). Couples who are infertile experience negative consequences including social stigmatization, ostracization, isolation, loss of marital stability, domestic violence, among others (3). In addition, infertility and/or delayed conception is often associated with psychological distress, (including depression and anxiety) (4) as well as poor quality of life (5). Across the world, women disproportionately affected by these negative outcomes compared to men (5–7) and tend to be blamed for infertility and/or delayed conception even when male causes of infertility and/or delayed conception are present (3, 8).

Despite the devastating consequences of infertility and/or delayed conception, access to high-quality fertility care is limited in many countries in low– and middle-income countries (9, 10). Disparities exist in terms of how people participate in and take up fertility care services, particularly diagnosis and treatment. Findings from a large systematic review showed that globally, only about half of all couples with infertility seek any form of infertility services (11). However, this is a global average and precise proportions of care seeking among those with infertility can vary from country to country (12).Several factors contribute to low access, spanning supply and demand. On the supply side, national health policies and benefit packages have often under-prioritized infertility, limiting availability, coverage, and public financing. (13–16). Additionally, inclusion of infertility in financial reimbursement and national insurance benefit packages is suboptimal in many low– and middle-income countries (13–15). Even in high income settings, subsidization of infertility is uneven (13, 17). Another factor that contributes to low access is sparse geographical availability of health facilities offering fertility care, that affects access of care due to logistical difficulties of traversing long distances (18). In terms of demand factors, several factors play a role. Lack of awareness around male and female causes of infertility, which is prevalent in many countries (19, 20) can affect uptake of services, as many people may not know that infertility can be resolved (9). In addition, social stigmatization of infertility often creates secrecy and inhibits care seeking (21). Furthermore, high costs of diagnosis and treatment of infertility negatively influence care seeking cost because of low affordability of fertility care, particularly in settings that do not have fully funded fertility care (22–24). In sum, global literature suggests that several factors located at individual, social or structural levels affect access to fertility care. From a policy standpoint, infertility care sits squarely within Universal Health Coverage (UHC) and reproductive rights. In many LMICs, fertility services remain minimally covered, compounding the cost, stigma, and access barriers described above. Framing delayed-conception care within UHC underscores the need for timely, affordable, evidence-based services, a context for the present study. Notably, the World Health Organization (WHO) 2023 report highlighted the substantial, globally distributed burden of infertility, reinforcing the urgency of system-level responses and equitable coverage.

In India, an estimated 3.9 to 16.8% of couples have infertility (3, 25), yet little research has been conducted on fertility care seeking. Most of the research on infertility in India has been conducted on the psychosocial experiences and consequences of infertility (26–29), as well as biomedical causes and treatment of infertility (30, 31). Social science findings consistently show a negative impact of infertility on social lives of those affected, which is similar global picture presented in the preceding paragraphs, while biomedical studies have demonstrated that male and female factors play a role in infertility. Despite studies from India indicating the perverse nature of infertility and its negative consequences, there remains a critical evidence gap on how, where, and under what conditions women seek care for delayed conception in routine settings.

Understanding care-seeking is important to inform the design of appropriate intervention that can assist couples and individuals resolve their infertility or delay in conception. Care-seeking is both a coping pathway and the first step toward timely diagnosis and treatment of infertility and/or delay in conception(32)(33). Unpacking the determinants of care seeking is equally relevant. Understanding influences of care seeking can be used to augment factors that promote care seeking, while mitigating those that prevent it. Given the foregoing, we conducted a study to document care seeking and its determinants among women who fail to get pregnant in Northern India.

## Materials and methods

### Study design

This was a mixed methods cross-sectional study which addressed the following research questions: 1) What are the characteristics of women who experience delay in conception? 2) What is the quality of life and mental health of women who experience delay in conception? and 3) What were the experiences and actions taken among women who experience delay in conception? The protocol for this study can be readily accessed in a separate publication (34). The results of the first two research questions have been reported in separate manuscripts where we describe the mental health status (35), quality of life (36) and qualitative experiences (37) of our study sample. In this manuscript, we focus on the issue of actions that women take, to describe the care seeking behaviors and its determinants among the study sample. This manuscript adheres to STROBE guidelines (38) for reporting quantitative observational studies (**Annex 1**).

#### Operational definition

Delayed conception (study definition): Women in the WINGS cohort who remained non-pregnant throughout 18 months of prospective follow-up despite being sexually active and not using contraception were classified as having delayed conception. To preserve study separation, eligible women were enrolled ≥14 days after exiting WINGS.

Clinical infertility (context definition): Per WHO, infertility is the failure to achieve a clinical pregnancy after 12 months of regular, unprotected sexual intercourse.

### Study population

The study population comprised women of reproductive age who completed 18 months without getting pregnant while enrolled in a preconception study known as the Women and Infants Integrated Interventions for Growth Study (WINGS) study (39). WINGS was a factorial, individually randomized trial conducted in urban Delhi to test the impact of an intervention package that consisted of nutrition, health, water, sanitation, and hygiene (WASH) and psychosocial care interventions among women of reproductive age. The women participants in WINGS trial were 18-30 years old, married, living with husband having no or one child and intend to have another child in next 1 year. The purpose of WINGS was to enable women of reproductive age to enter pregnancy in good health, free of sexually transmitted infections (STIs), and well-nourished. WINGS was implemented in the low to mid-socioeconomic neighborhoods of urban Delhi, India as detailed in the study protocol (34). These neighborhoods were chosen based on the potential for demonstrating impact of interventions provided before and during pregnancy, based on evidence that poor health prior to conception (pre– and peri-conception period) is linked to birth outcomes (40). To participate in the present study, women were recruited after they exited from WINGS without getting pregnant. Hence, the eligibility was delayed conception defined as The minimum sample size needed to estimate infertility with 2% precision and 95% CI from this sample would be 1223, assuming a prevalence of 17% as reported in the literature and a finite population of 12,500.(41) An allowance of 25% for rejection/ incomplete nonresponses was made and the final sample size was 1530.

### Data collection procedure

All married women who had completed 18 months of participation in the WINGS project without getting pregnant were approached by trained field workers. These women were contacted at least 14 days after their exit from the WINGS program, as this was a separate study.

Women expressing interest in participating were invited for an appointment, during which research assistants, who were professionally trained nurses, provided comprehensive information about the purpose, procedures, potential risks, and benefits of this study. Informed consent was obtained following a thorough explanation, with particular emphasis on the voluntary nature of participation and assurances of confidentiality. Research assistants verified participants’ understanding before enrolment, reiterating that declining participation would have no negative consequences. Written informed consent was obtained from all participants.

A total of 1,530 married women consented and completed the survey. Subsequently, the survey questionnaire, consisting of a semi-structured questionnaire (Module 1 and 2) [**Supplementary File 1**], was administered face-to-face by the research team. The first module collected information on women’s sociodemographic characteristics, including age, marital status, husband’s age, and duration of marriage. Data on fertility history and intentions were gathered, such as history of childbirth, age at first childbirth, date of last delivery, intended number of children, presence of adopted or fostered children, and whether the husband had fathered children. Information on pregnancy attempts and behavior was also collected, duration of trying to conceive, frequency of unprotected intercourse, and women’s perceptions of delays in conception. Additionally, the survey captured perceived stigma and emotional experiences, including women’s and their husbands’ views on delayed conception and feelings of isolation or discrimination. In the second module, information was collected on the actions undertaken by women in response to their delayed conception. This included their knowledge of available sources of care, preferred types of healthcare providers (such as general practitioners, specialist doctors, traditional doctors, and traditional healers like priests), and the use of alternative forms of care, including BHMS (Bachelor of Homeopathic Medicine and Surgery), Ayurveda, and other indigenous systems. Additionally, the module captured reasons for not seeking care and explored the role of the partner in supporting or facilitating care-seeking behaviour and its reasons.

### Outcomes

The primary outcome of this study was whether care was sought for delayed pregnancy or not and was assessed as a categorical variable. The other outcomes were sources of care sought and role of husbands in care-seeking.

#### Exposure variables

The exposure variables included social demographic characteristics of the married women and their families (such as age, education, occupation, religion, duration of marriage, wealth index etc.) fertility intentions of women, substance use, and medical and sexual history of women. The selection and adaptation of these variables were informed by previous research and surveys related to delayed conception (35, 36).

Formal (Appropriate) sources for care-seeking were defined as when women sought care for delayed conception from rregistered facilities such as government hospitals, community health centers, private clinics, or IVF centers or Licensed paramedical staff (e.g., nurses, ANMs, or lab technicians) when providing fertility-related medical services under clinical supervision, or helpline or non-governmental organization, or Government social services. Hence, formal health-care providers in our study are those legally trained, accredited, and integrated within India’s biomedical health system, offering diagnosis or treatment for infertility in accordance with recognized medical standards.

Informal-sources for care-seeking were defined as when women sought care for delayed conception from unregulated or traditional sources of advice or treatment such as traditional healers, herbalists, religious leaders, or parents or family elders/ members, or friends, or husband, or peer-support group, or research study staff,r etc.

### Data analysis

Data were first checked for completeness, cleaned, and subsequently analysed using STATA version 16.0 (Stata Corp). Descriptive statistics were computed to summarize sample characteristics, with continuous variables reported as means and standard deviations, and categorical variables presented as proportions. To assess household economic status, a wealth index was constructed based on asset ownership and principal components analysis (PCA) was employed to generate a continuous scale reflecting relative household wealth. The resulting total scores were used to categorize the population into five wealth quintiles: the poorest, very poor, poor, less poor, and the least poor.(35)The proportion of women who sought care for delayed conception, as well as the sources of care utilized, was calculated. Reasons for not seeking care, reported barriers to care-seeking, and additional actions taken by women, such as modifying sexual behaviour, knowledge of service availability, and support received from husbands, will be analysed by calculating frequencies and percentages.

Univariable logistic regression was performed to explore associations between each independent variable, such as women’s sociodemographic and family characteristics, fertility intentions, wealth index, substance use, medical and obstetric history, and sexual history—and the primary outcome, i.e., care-seeking for delayed conception. For each univariable analysis, the corresponding p-value was recorded. Variables were then categorized into two sets based on their p-values: those with p<0.20 and those with p≥0.20. All variables with a p-value less than 0.20, along with clinically and biologically important factors, were considered for inclusion in the multivariable logistic regression model. A stepwise backward elimination approach was used for multivariable modelling. Variables with a p-value ≥0.05 were sequentially removed, and the model was re-estimated until only statistically significant variables remained. Subsequently, variables initially excluded (p≥0.20 in univariable or ≥0.05 in multivariable analysis) were individually reintroduced into the final model to reassess their significance. Any variables found to be statistically significant at p<0.05 were retained in the final model. Diagnostic tests for model assumptions, including linearity, heteroskedasticity, multicollinearity, normality of errors, and independence of residuals, were conducted to ensure the robustness of the final regression model.Model fit was assessed using the Akaike Information Criterion (AIC), and Bayesian Information Criterion (BIC) with the final model demonstrating optimal fit based on lower AIC/BIC value. (42)

### Ethical approval

This study, which involved human participants, received ethical approval prior to its initiation. The study protocol was first reviewed and approved by the Ethics Review Committee of the Society for Applied Studies (SAS), the institution of the principal investigator (approval number: SAS/ERC/RHR-Infertility/2020; approval date: 9 January 2020). Additional approval was obtained from the World Health Organization (WHO) Ethical Review Committee (approval number: A-ID A65998; approval date: 13 February 2020). Informed written consent was obtained from all participants before enrollment. Detailed information regarding the study’s objectives, potential risks, and confidentiality protections were provided to each participant. Considering the sensitive nature of the topic, trained nurse research assistants ensured that participants fully understood the study details and that their participation was entirely voluntary, without any pressure or coercion.

## Results

A total of 1,530 women were included in the study, with a mean age of 26.8 years (SD 3.3). Nearly half (48.8%) had one living child, and among these, the mean age of the youngest child was 47.1 months (SD 29.7). Most participants lived in extended or joint family households (56.8%), while 43.2% resided in nuclear families. The average household size was 4.8 members (SD 2.7).

The mean educational attainment among women was 10.2 years of schooling (SD 4.3). Most women identified as housewives, and only 6.0% were employed outside the home. The mean age of husbands was 28.6 years (SD 3.9), with a mean of 11.0 years of schooling (SD 3.8). Most husbands were employed in the private sector (71.5%) and 4.2 % were unemployed. Over three-quarters (76.5%) reported home ownership, with ownership most often in the name of a male member (82.4%). Almost all households had at least one bank or post office account (90.5%), but only 11.8% reported having health insurance or coverage under a health scheme. The mean annual family income was USD 2,801.21 (SD 1,405.5) and households holding a below-poverty-line card comprised 4.8% of the sample. (**Table 1)**

**Table 1:**
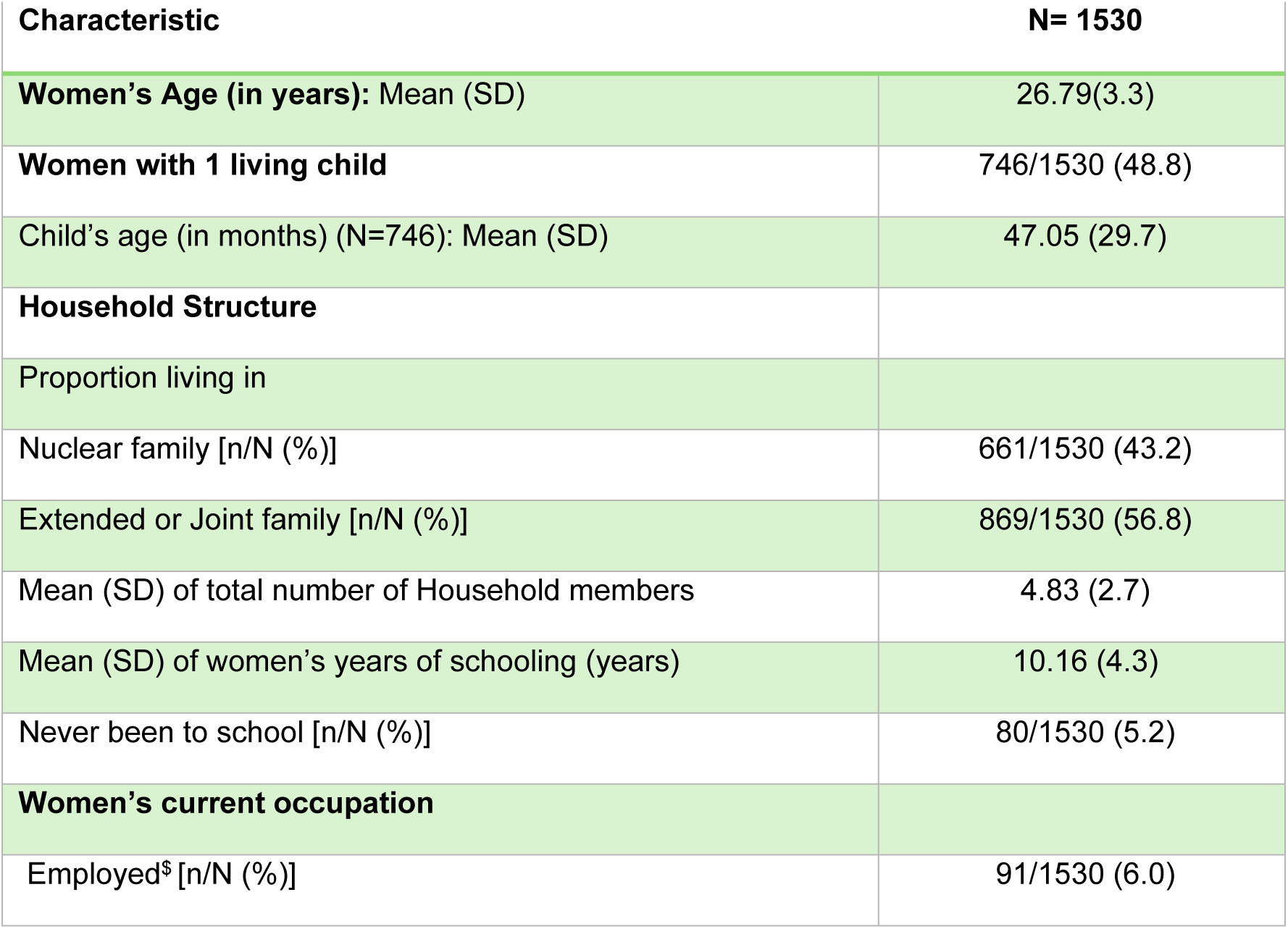
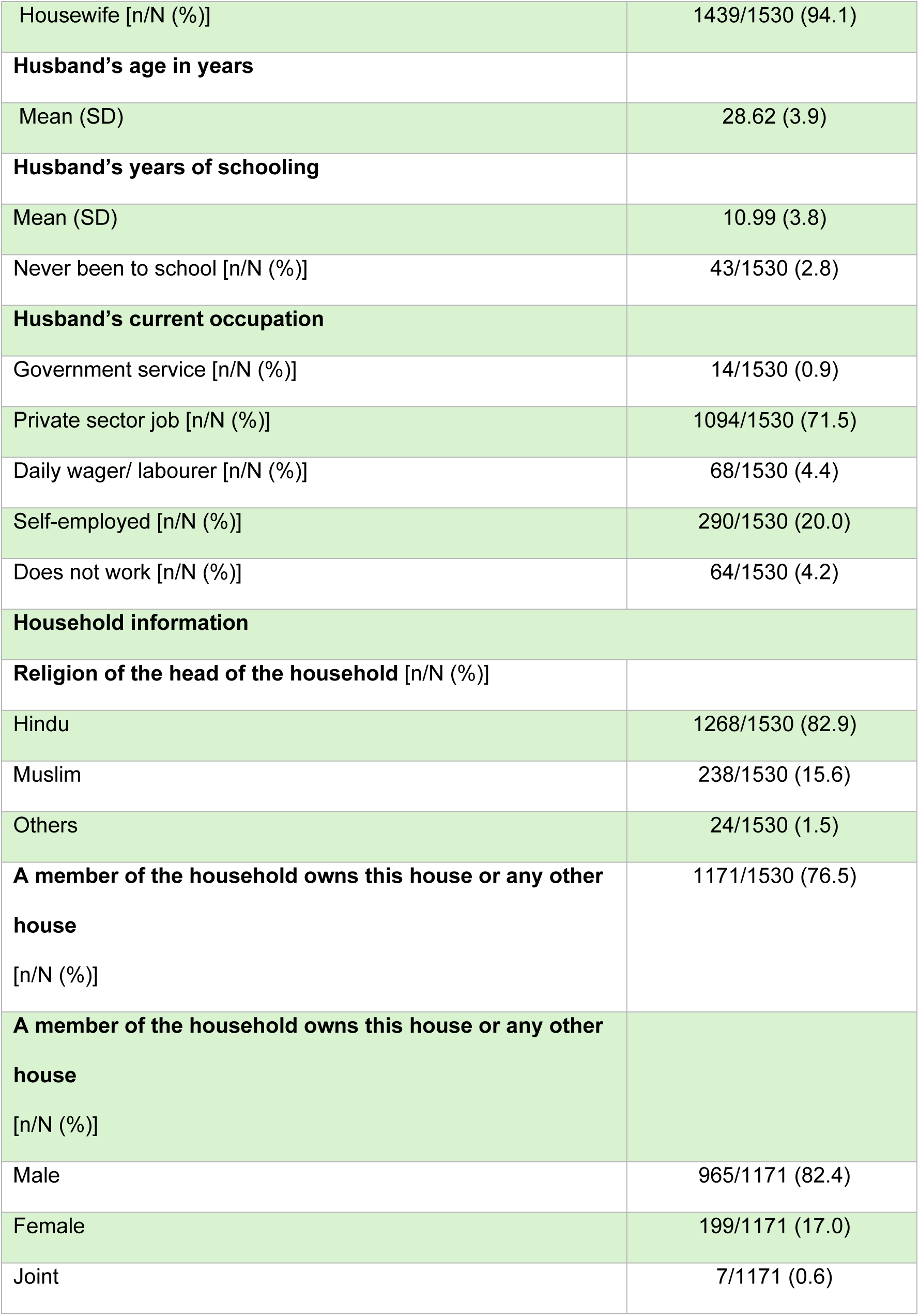
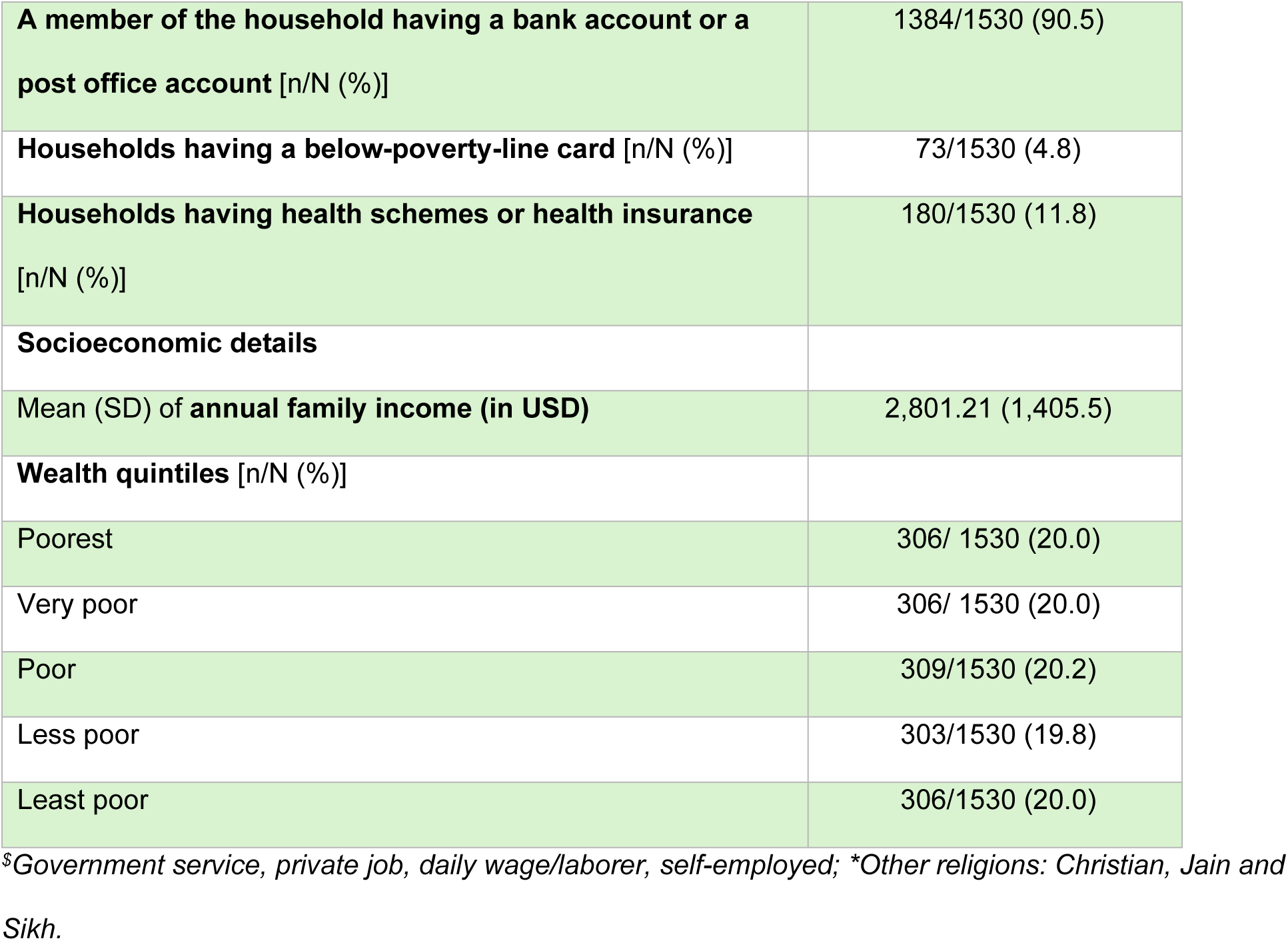
Baseline characteristics of the married women included in the study.

**S1 Table** shows among 1,530 women with delayed conception, about 70% sought help, most commonly from both formal and informal providers. Specialist doctors were the main source of treatment, while traditional healers, family, and religious leaders also played major roles, offering mostly advice, counselling, or herbal remedies rather than biomedical interventions.

Among 1,530 women, nearly half (49.4%) had never had a biological child, and 3.1% reported having adopted, foster, or stepchildren. The mean age at first birth was 22.2 years (SD 2.7), and women had been married to their current husband for an average of 6.4 years (SD 3.2). In 40.9% of cases, the partner had fathered a child with another woman. Prior to enrolment, 80.0% reported intending to become pregnant. The average duration of attempting to conceive was 3.2 years (SD 2.1). Most women (50.4%) reported unprotected sexual intercourse more than twice a week, while 28.3% reported twice weekly, and smaller proportions reported less frequent intercourse**. (Table 2)**

**Table 2:** Reproductive history and fertility intentions of the women.

| Variables | N=1530 |
| --- | --- |
| **Women who never had a child [n/N (%)]** | 755/1530 (49.4) |
| **Women who have any adopted, fostered, or stepchildren [n/N (%)]** | 47/1530 (3.1) |
| **Maternal age in years at first birth (Mean, SD)** | 22.15 (2.7) |
| **Years woman has been married to her current husband (Mean, SD)** | 6.42 (3.2) |
| **Partner fathered a child** (from another woman) | 626/1530 (40.9) |
| Intended pregnancy before enrolment in the primary trial* | 1224/1530 (80.0) |
| **Duration of trying to conceive** (in years) (Mean, SD) | 3.21 (2.1) |
| **Unprotected sexual intercourse** |  |
| Once every month | 27/1530 (1.8) |
| Twice a month | 100/1530 (6.5) |
| Once a week | 199/1530 (13.0) |
| Twice a week | 433/1530 (28.3) |
| More than twice a week | 771/1530 (50.4) |
| **Reports delay in pregnancy** (as per Woman) | 1374/1530 (89.8) |
| **Reports delay in pregnancy** (as per husband) | 763/1530 (49.9) |
| **Experienced from partner** |  |
| Physical abuse | 62/1530 (4.1) |
| Emotional abuse | 262/1530 (17.1) |
| Verbal abuse | 131/1530 (8.6) |
| Denial of financial support | 34/1530 (2.2) |
| Divorce | 30/1530 (2.0) |
| Abandonment | 31/1530 (2.0) |
| **Experienced from anyone else in the family for taking too long to become pregnant** |  |
| Physical abuse | 47/1530 (3.1) |
| Emotional abuse | 748/1530 (48.9) |
| Verbal abuse | 279/1530 (18.2) |
| Denial of financial support | 51/1530 (3.3) |
| Divorce | 41/1530 (2.7) |
| Abandonment | 51/1530 (3.3) |

*\*The participants of this study were earlier a part of trial names as WINGs.*

A large majority (89.8%) perceived a delay in pregnancy themselves, and half (49.9%) reported that their husband also perceived a delay. Experiences of intimate partner violence were reported by 4.1% (physical abuse), 17.1% (emotional abuse), and 8.6% (verbal abuse), with smaller proportions reporting denial of financial support (2.2%), divorce (2.0%), or abandonment (2.0%). Experiences of abuse from other family members due to delayed pregnancy were also notable, including emotional abuse (48.9%), verbal abuse (18.2%), and physical abuse (3.1%). **(Table 2)**

Of the 1,530 participants, 69.9% (n=1,069) sought care for delayed conception, while 30.2% (n=461) did not seek care. Among those who sought care, 8.4% (n=129) accessed only formal (appropriate sources), 1.7% (n=26) used only non-appropriate sources, and the majority (59.7%, n=914) sought care from both appropriate and non-appropriate sources. (**Table 3)**

**Table 3:** Distribution of care-seeking and its sources among all participants.

| Category | n<br>(N=1530) | % |
| --- | --- | --- |
| Did not seek care | 461/1530 | 30.1 |
| sought care | 1069/1530 | 69.9 |
| Sought care from only formal source | 129/1069 | 12 |
| Sought care from only informal sources | 26/1069 | 2.4 |
| Sought care from both formal and informal sources<br>(mixed) | 914/1069 | 85.5 |

Of the total participants, 30.1% (n = 461) women did not seek help for delayed conception. The most common reasons reported for not seeking care were the belief that they did not have a problem (69.9%), hope that the problem would resolve on its own (76.4%), and attempting to solve the issue independently (38.7%). Financial barriers were also frequently cited, with 37.8% having heard that treatment is very expensive and 33.6% stating they did not have the money. Additional barriers included lack of contacts to access professional care (27.1%), difficulties with transport (24.2%), embarrassment or shame (16.2%), and concerns about social perceptions, including what family (15.1%) and friends (5.6%) might think. Other reported reasons were concerns about treatment side effects, previous negative experiences with providers, and lack of awareness about available fertility treatments. (**Table 4)**

**Table 4:** Reasons for not seeking care among the women.

| Proportion of women who did not sought help | 461 (30.1) |
| --- | --- |
|  | **n = 461** |
| **Reasons for not seeking help** |  |
| Didn't perceive they had a problem | 320 (69.87) |
| Not aware of existence or possibility of fertility treatment | 22 (4.80) |
| Concerned about what my friends might think | 26 (5.64) |
| Concerned what husband might think | 34 (7.42) |
| Did not have any access to professional care | 124 (27.07) |
| Solving the problem by self | 177 (38.65) |
| Concern that they might be seen as physically weak for having delayed conception | 36 (7.86) |
| Problem with transport or travelling to appointments | 111 (24.24) |
| Believe that this problem would get better by itself, and I would get pregnant | 350 (76.42) |
| Concerned about family's opinion | 69 (15.07) |
| Felt embarrassed or ashamed | 74 (16.16) |
| Preferred alternative forms of care | 56 (12.23) |
| Unable to afford the financial costs involved | 154 (33.62) |
| Concerned that women might be seen as not valuable | 19 (4.15) |
| Thought that professional care probably would not help | 21 (4.59) |
| Too unwell to ask for help | 49 (10.70) |
| Concern that people might find out about woman's issue of delay in conceiving. | 27 (5.90) |
| Dislike talking about her feelings, emotions or thoughts | 132 (28.82) |
| I was concerned that people might not take me seriously if they found out I was having delay in conception. |  |
| Concerned about the treatments available (e.g. medication side effects) | 50 (10.92) |
| Had previous bad experiences with fertility experts or other such health care providers | 31 (6.77) |
| Heard treatment is very expensive | 173 (37.77) |
| Did not have the money | 153 (33.41) |
| Heard these treatments are for long duration and not effective | 76 (16.59) |
| Awareness among woman that problem is with my husband, and he will not seek care | 20 (4.37) |

The screen plot **(S 1 Fig.**) illustrates the distribution of eigenvalues across extracted factors. There is a marked decline in eigenvalues after the first factor, with a noticeable “elbow” at the third factor, after which the curve flattens. According to the Kaiser criterion (eigenvalues > 1) and the shape of the scree plot, a three-factor solution is appropriate. This indicates that three main underlying dimensions best summarize the patterns of barriers to delayed conception’s care-seeking in this population

Factor analysis was conducted on binary-coded reasons for not seeking care for delay in conception among women who did not seek help (n=194); (**Table 5**). The screen plot and eigen value criteria supported the retention of three underlying factors, which together explained a substantial proportion of the total variance. The first factor (social stigma and interpersonal barriers) captured concerns about friends’ and husband’s opinions, fear of being seen as weak, secrecy, shame, and lack of awareness, highlighting the powerful role of social norms and stigma in shaping help-seeking. The second factor (financial and structural barriers) included high loadings for lack of money, perceived high costs, and hearing that treatment was expensive or lengthy, reflecting economic and health-system challenges. The third factor (self-reliance and optimism) encompassed beliefs that the problem would resolve on its own, a preference for self-management, and dislike of discussing personal issues. (**Table 5**).

**Table 5:**
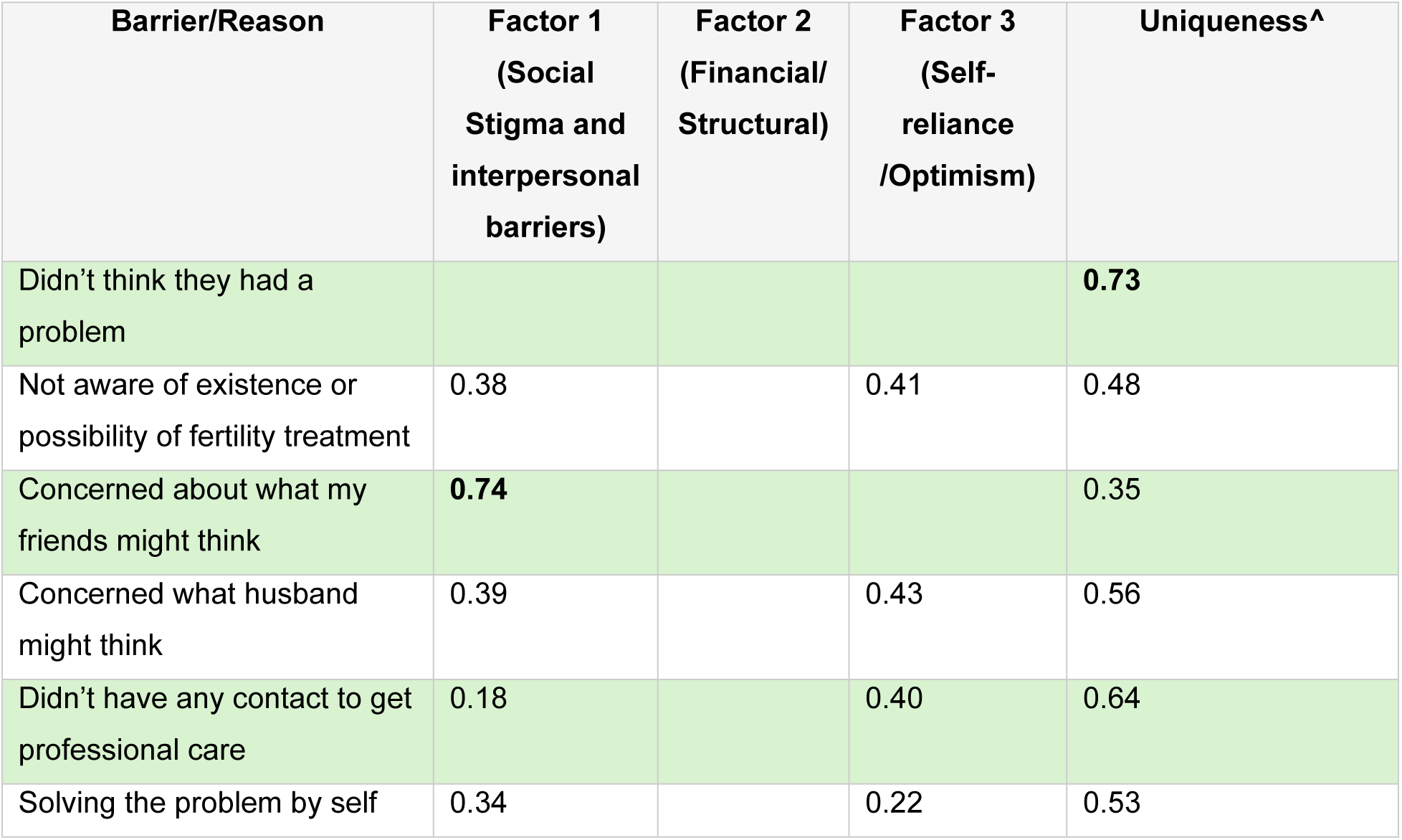
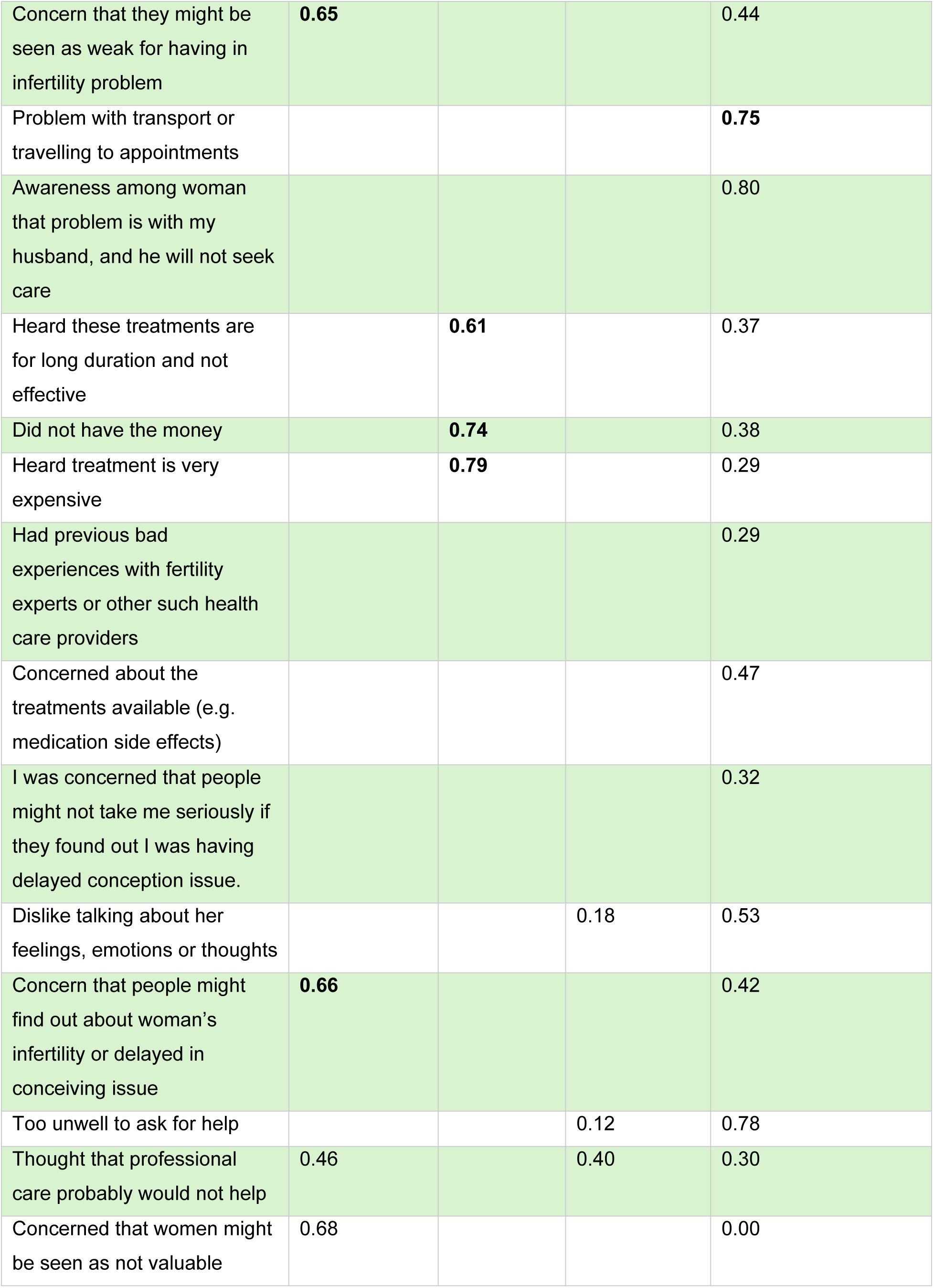
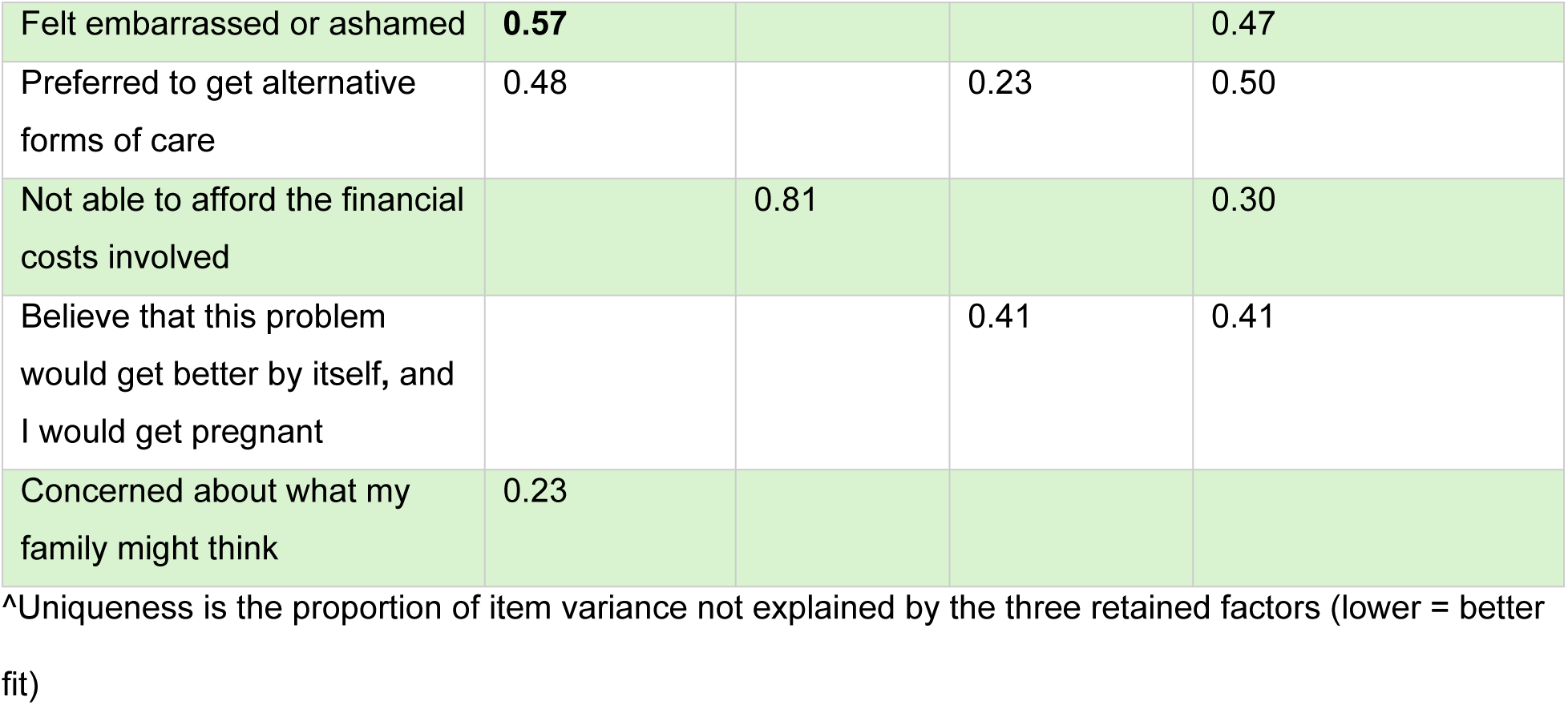
Factor analysis for reasons for not seeking care.

Several reasons, such as lack of awareness and preference for self-management, showed moderate cross-loadings on multiple factors, indicating that many barriers to care-seeking are interrelated and reflect both social and structural influences. Overall, these findings suggest that interventions to improve care-seeking for delay in conception must address not only health system factors and affordability but also social stigma, psychological attitudes, and informational gaps. (**Table 5**).

Multivariable logistic regression identified several independent predictors of care-seeking among women with delay in conception (**Table 6**). Each additional year spent trying to conceive was associated with a 60% increase in the odds of seeking care (aOR: 1.6, 95% CI: 1.4–1.7, p < 0.001). Women who perceived that it was taking longer than expected to conceive had more than three times the odds of seeking care compared to those who did not (aOR: 3.1, 95% CI: 2.1–4.5, p < 0.001). Similarly, women who reported feeling isolated had 1.7 times higher odds of seeking care (aOR: 1.7, 95% CI: 1.2–2.4, p < 0.001). Additionally, older age groups showed markedly lower odds of seeking care: women aged ≥25 to <35 years had reduced odds compared with those aged 18–24 years (aOR: 0.6, 95% CI: 0.5–0.8, p = 0.001), and women aged ≥35 years had the lowest odds (aOR: 0.2, 95% CI: 0.1–0.5, p = 0.002). Experiencing emotional abuse from family members (aOR: 1.6, 95% CI: 1.2–2.2, p = 0.001) and from husbands (aOR: 1.7, 95% CI: 1.2–2.4, p = 0.005) were also significantly associated with increased odds of care-seeking. Women reporting heavy menstrual bleeding had 1.5 times higher odds of seeking care compared to those who did not (aOR: 1.5, 95% CI: 1.0–2.2, p = 0.005). In contrast, having regular monthly periods was associated with a 40% reduction in the odds of care-seeking (aOR: 0.6, 95% CI: 0.4–0.9, p = 0.03).

**Table 6:** Multivariable model representing the determinants of care-seeking for delayed conception among study participants.

| Variables | Sought Care |  |
| --- | --- | --- |
|  | Un-adjusted OR (95% CI), (p-value)<br>(Source: All types, including Formal, Informal and Mixed) | Adjusted OR (95% CI), (p-value)<br>(Source: All types, including Formal, Informal and Mixed) |
| **DEMOGRAPHIC** |  |  |
| Women age |  |  |
| >= 18 to 21 years | Reference | Reference |
| 22 to 27 years | 0.8(0.6, 1.0) (p=0.07*) | 0.6 (0.5, 0.8) (p=0.001) |
| 28 years and above | 0.6 (0.2, 1.4) (p=0.24) * | 0.2 (.1,0.5) (p=0.002) |
| **FERTILITY INTENTIONS** |  |  |
| Total number of years trying to conceive | 1.7 (1.6,1.9) (p=0.000*) | 1.6 (1.4,1.7) (p=<0.001***) |
| Perception of the woman that conception is taking longer |  |  |
| Yes | 5.4(3.8,7.7) (p=0.00*) | 3.1 (2.1,4.5) (p=<0.001***) |
| No | Reference | Reference |
| Women felt isolated |  |  |
| Yes | 3.2(2.5,4.0) (p=0.000***) | 1.7 (1.2,2.4) (p=<0.001***) |
| No | Reference | Reference |
| Women verbally abused by partner/husband |  |  |
| Yes | 1.2 (0.8,1.9) (p=0.3) |  |
| No | Reference |  |
| Women emotionally abused by family members |  |  |
| Yes | 2.3** (1.8,2.9) (p=0.000) ** | 1.6 (1.2,2.2) (p=0.001*) |
| No | Reference | Reference |
| Women emotionally abused by husband |  |  |
| Yes | 1.0 (0.7,1.3) (p=0.9) | 1.7 (1.2,2.4) (p=0.005) |
| No | Reference |  |
| Women verbally abused by family members |  |  |
| Yes | 1.3 (1.0,1.8) (p=0.10) |  |
| No | Reference |  |
| **MEDICAL HISTORY** |  |  |
| Regular period every month |  |  |
| Yes | 0.6*(0.4,0.9) (p=0.004) | 0.6* (0.4,0.9) (p=0.03*) |
| No | Reference | Reference |
| Bleeding Heavy during periods |  |  |
| Yes | 1.4 (1.0,2.0) (p=0.03) * | 1.5(1.0,2.2) (p=0.005) |
| No | Reference |  |
| Bleeding after sexual intercourse |  |  |
| Yes | 0.6 (0.3,1.2) (p=0.2) |  |
| No | Reference |  |

Additionally, exploratory models assessing interactions revealed that the association between regular monthly periods and care-seeking was significantly modified by women’s perception of social isolation (interaction aOR: 3.78, 95% CI: 1.20–11.89, p = 0.023). While regular cycles alone were associated with reduced odds of care-seeking, women who also reported feeling isolated had markedly higher odds of seeking care, suggesting that social context may override biological reassurance. No significant interactions were observed between regular periods and emotional abuse (from husband or family) or age group, indicating that the protective effect of regular cycles on care-seeking is largely independent of these factors. We also explored whether the association between years of trying to conceive and care-seeking was modified by age group; interaction terms were not statistically significant, suggesting that the effect of duration of delay in conception on care-seeking was consistent across age categories. A three-way interaction involving regular menstrual cycles, age, and years trying to conceive could not be reliably interpreted due to sparse data in some subgroups.

**S2 Table** presents the multivariable model showing determinants of care-seeking for delayed conception from all sources versus formal and mixed sources of care among study participants. Women aged 25 years and above were significantly less likely to seek care for delayed conception compared to those aged 18–24 years. The likelihood of seeking care increased with the perception that conception was taking longer than expected (aOR = 1.6, 95% CI: 1.4–1.7, p < 0.001) and was even higher when seeking care from formal or mixed sources (aOR = 4.6, 95% CI: 3.2–6.6, p < 0.001). Psychosocial factors such as feeling isolated (aOR = 1.7, 95% CI: 1.2–2.3) and experiencing emotional abuse from partners (aOR = 1.6, 95% CI: 1.2–2.4) or family members (aOR = 1.5, 95% CI: 1.2–1.8) were also independently associated with care-seeking. Women reporting heavy menstrual bleeding were more likely to seek care (aOR = 1.5, 95% CI: 1.0–2.2, p = 0.03), whereas those with regular monthly periods had significantly lower odds of seeking care (aOR = 0.7, 95% CI: 0.4–0.9, p = 0.02). Similar patterns were observed when restricted to women seeking care from formal and mixed sources, indicating that both psychosocial and reproductive health factors were important determinants of fertility-related care-seeking behavior.

## Discussion

This study provides important information on the prevalence and determinants of care-seeking among women with delayed conception in low-to middle income setting of Northern India. Findings show that nearly 70% of women sought care for delayed conception. Of those that sought care, exclusive care-seeking from formal sources was low (8.4%), moderate proportions exclusively accessed informal sources (12%), while a clear majority accessed mixed sources (85%). Multivariable analysis found that a longer duration of trying to conceive, perceiving the process as taking too long, feeling isolated, experiencing emotional abuse from husbands or family, and heavy menstrual bleeding were independently associated with increased odds of care-seeking. Conversely, having regular monthly periods was associated with lower odds of seeking care. Several aspects of these findings are important and warrant further discussion.

First, the high care-seeking rate is particularly striking, compared to other similar settings, which report lower levels of care seeking in different settings such as China (43, 44), sub-Saharan Africa (45, 46), and indeed globally (11). Studies have consistently shown that people with infertility and/or delay in conception endure it in silence, in secrecy, isolation and self-stigmatization (47). Our finding of a relatively larger proportion seeking care may be because nearly half of these women had never had a biological child, which is particularly important in terms of the impact of delayed conception on an individual. Other analyses have shown that having at least one child, or higher parity, is likely to decrease care-seeking, at both conceptual (48) and empirical (12) levels. The high level of care seeking in our study is notable given that, formal sources (on its own or in combination with informal care seeking) were sought by 68% of women. A possible explanation could also be related to the fact that most women in our sample had previously participated in a pre-conception RCT (34), which may have sensitized them on issues related to health care seeking.

Despite the high level of care-seeking, barriers persisted for many women, and about one-third of them did not seek any help, due to a mix of individual, social and structural barriers. At the individual levels failure to seek care was most often due to perceptions that they didn’t have a problem, or the problem would resolve on its own, lack of perceived severity, dislike of talking about their own situation, or a preference for self-management. Social and psychosocial barriers such as embarrassment, shame or concerns about what family members might think, stigma and fear of side-effects were common. Structural factors related to financial and logistical barriers, such as perceived cost, lack of contacts or referrals to professional care, and transport difficulties, were common. Such factors are highlighted by other studies in similar settings (31, 45).

Second, our study highlights independent predictors of care-seeking. Each additional year spent trying to conceive was associated with a 54% increase in the odds of seeking care. This is expected given that time is an important construct in delayed conception, and the duration of delay in conception which can affect how an individual copes with this problem (49), or prompt actions to try and resolve it (50). Independent of the duration of delay in conception, women who perceived that it was taking too long to conceive had nearly three times the odds of seeking care compared to those who did not. The association of longer duration of trying to conceive and greater subjective concern (“felt it was taking too long”) with increased care-seeking is intuitive and aligns with a threshold effect, where sustained failure to conceive triggers a shift from passive waiting to active help-seeking, as indicated in other studies (30). Other studies have highlighted how perceived infertility or delay in conception, regardless of actual waiting time to pregnancy, can affect care seeking (51). In our study setting, the expectation is that childbirth would occur within one year or marriage, and the passage of time longer than a year tends to trigger a perception of delay. All the participants in our study had waited for at least 18 months to conceive without success. While it is important for people to know that time to pregnancy can vary (51), the duration of 18 months of waiting could be expected to trigger help seeking.

An important group of independent predictors relate to the psychosocial factors: experiencing isolation, emotional abuse from husband or emotional abuse from family members were all independently associated with higher odds of seeking care. The impact of the social environment is important given that almost half of women reported emotional abuse from their families and husbands due to delayed conception, and a considerable proportion experienced emotional or verbal abuse from their partners. Our study highlights psychosocial environments as being a major determinant of care seeking (30). The mechanism through which this occurs is unclear, but it is possible that women seek help in order to stop emotional abuse from their spouses and families. This may reflect a drive to resolve infertility and or delayed conception-related stigma or distress, or possibly greater familial/social pressure to “act” when distress becomes visible, as suggested by other researchers (30). This is particularly relevant given that India is a highly pro-natal society where childbearing is valued by the society and the failure to bear a child is discouraged and punished for example through stigma, isolation or discrimination. At the same time, emotional abuse is detrimental to quality of life (36) and can exacerbate mental distress (35) should therefore be mitigated.

Our study showed that women who experienced heavy menstrual bleeding also had higher odds of seeking care, while having regular monthly periods was associated with lower odds of seeking care. heavy menstrual bleeding which could elicit subjective perceptions of poor health; it is possible that as predicted by help-seeking models, poor subjective health could influence care seeking (48). However, it is unclear how having regular monthly period reduces the odds of seeking care as it is a constant indicator and reminder of delayed conception and infertility. We hypothesize that women with regular menstrual cycles may perceive themselves as less likely to have an underlying problem, leading to delayed or deferred care-seeking, regular menses reinforce saliency of absence of pregnancy. This is supported by the observation that the most common reason given by women who did not seek help was that they didn’t think they had a problem. Our interaction analysis further showed that this protective effect of regular cycles was significantly modified by women’s perception of social isolation: while regular cycles alone were associated with reduced odds of seeking care, women with regular cycles who also reported feeling isolated had markedly higher odds of care-seeking. This suggests that the social context can override biological reassurance, prompting care-seeking even among women who otherwise view their cycles as normal. Final point to note in terms of predictors of care-seeking is that unlike other studies (12, 52) demographic factors such as age, education status and economic status were not independent predictors of care seeking; however, while our large sample size provides credible data, we acknowledge that such predictors could vary between geographies (12) and over time.The strong association between heavy menstrual bleeding and care-seeking likely reflects greater symptom burden, prompting medical consultation.

Third, apart from predictors of care seeking, our study provides an important insight into the predominant pattern of mixed source utilization that highlights the complexity of the delayed conception care landscape, where biomedical, traditional, and social care pathways often coexist rather than substitute for one another. This pattern likely reflects a combination of persistent sociocultural beliefs, widespread stigma around infertility or delayed conception, and limited confidence in formal health services. The pattern of utilization reflects similar patterns described in reviews and studies from resource-poor settings, where women often “shop” across multiple providers due to uncertainty, lack of clear guidance, or dissatisfaction with previous (45, 53–55). This has a significant impact on discontinuity of care, duplication or unnecessary treatments, diagnostic delays and multiplicity of treatments, and results in negative experiences and poor quality of care among those who are seeking care for delayed conception(54, 55).

### Implications for policy and practice

By providing population-based data on care-seeking, our study provides insights into the demand for fertility care, barriers to formal biomedical sources of care, and factors that can be modified to facilitate access to care, with direct implications for the design and delivery of infertility and/or delayed conception services in India. The tendency to seek care from both formal biomedical and informal sources, is important and indicates the need to ensure that all actors in care seeking journeys have adequate information about the nature of infertility and/or delayed conception, which interventions are effective, and where to seek care so that they can pass this information to. A significant proportion of participants in our study, as is the case in other countries, seek informal sources, who are easy to access and cheaper, but whose advice may not always be accurate, skill set may not be quality assured, and the treatments or medicine they provide may not always be safe. A qualitative study with a subset of our participants indicated that awareness is low and that herbal and other home remedies are common (50), suggesting that public education initiatives on infertility and/or delayed conception are needed to increase awareness communities in our study setting.

The multiplicity of factors that prevent care-seeking triggers suggests that interventions must be multifaceted to further enhance it. For example, it is important to address financial constraints through insurance coverage of infertility and/or delayed conception treatments, while educating the public to improve awareness regarding the benefits of timely, evidence-based interventions. At the same time health systems should integrate psychosocial counselling and other mental health interventions into infertility and/or delayed conception care, address stigma, and include couple-based and family-based interventions to mitigate emotional abuse directed at women with infertility and/or delayed conception. This will ensure that communities, families and spouses are educated to support rather than blame these women. Studies have shown that spousal and family support is protective in mitigating distress and stigma experienced by people with infertility (56, 57).

Importantly, our findings suggest that utilisation of the public health system was limited. There is a need for health system strengthening to expand infertility and/or delayed conception care coverage, especially in the public sector, and to include routine psychosocial screening and support. Expanding the reach and quality of public sector fertility services, alongside culturally sensitive information campaigns, may help to shift patterns to increased utilization of fertility care in public hospitals. National policies should prioritize infertility as an integral part of reproductive health and offer subsidized, accessible, and comprehensive care.

#### Implications for research

Gendered differences in infertility and/or delayed conception care-seeking have been reported elsewhere (51, 52) and we hypothesize that many male partners of the women that sought care in our study did not seek care. Qualitative findings with a few women from our sample indicated the hesitancy of husbands to seek care (58); our unit of analysis on care-seeking is therefore women and may not be generalized to both partners in a couple. Future studies are needed on care seeking with male partners. Given the high rates of care-seeking from mixed sources, further qualitative research is needed to understand motivations and barriers, and effects that these have on outcomes of people with infertility and/or delayed conception. Future research could also explore interventions that address both biomedical and psychosocial aspects of infertility care-seeking and rigorously evaluate strategies to shift care-seeking towards competent and effective providers.

### Strengths and Limitations

This study is among the largest and most detailed cross-sectional investigations of care-seeking among women with delayed conception in urban India, with a sample size of over 1,500 women, recruited from the community. This approach captures a wide range of clinical, demographic, social, and psychosocial determinants of care seeking. Other researchers have noted the importance of using non-clinic-based samples to assess infertility care-seeking (12) given that a substantial portion of people with infertility and/or delayed conception never seek care from formal health care services (11). Our study quantifies both formal and non-informal sources of care, including mixed-pathway utilization, providing useful information that can inform strategies to improve care-seeking. Rigorous data collection, using a structured survey and careful operational definitions, enhances the validity and reliability of our findings and their generalizability to other urbanized low-to-middle socio-economic Indian settings.

Several limitations should be considered. First, as with most cross-sectional studies, causal inferences cannot be made. The study population, drawn from women who participated in a trial with a preconception component and may have higher health awareness and may not be representative of all women experiencing delayed conception in India. Additionally, women belong to a particular neighbourhood of a North Indian city which also limits generalizability. The classification of care sources relied on self-report and may be subject to recall or misclassification bias. Our study did not assess the quality, continuity, or content of care received from different sources, nor did it include perspectives from male partners for quantitative data generation, which are increasingly recognized as important in infertility and/or delayed conception care dynamics. Future studies are needed on care seeking with male partners.

## Conclusion

This study shows that while most women with delayed conception in urban India sought care, the majority relied on fragmented pathways combining formal and informal providers. Care-seeking was shaped not only by biomedical concerns but also by psychosocial and social factors, including perceived delays, isolation, and emotional abuse. At the same time, financial, structural, and stigma-related barriers prevented many from accessing care. These findings underscore the need for integrated, equity-focused infertility and/or delayed conception services that combine medical care with psychosocial support, reduce stigma, ensure financial protection, promote fertility education, and expand access through strengthened public health systems.

## Author contributions

**Conceptualization:** Sarmila Mazumder, Barsha Gadapani Pathak, and Gitau Mburu

**Data curation:** Barsha Gadapani Pathak

**Formal analysis:** Barsha Gadapani Pathak.

**Funding acquisition:** Sarmila Mazumder, and Barsha Gadapani Pathak

**Investigation:** Sarmila Mazumder, and Barsha Gadapani Pathak

**Methodology:** Sarmila Mazumder, and Barsha Gadapani Pathak

**Project administration:** Sarmila Mazumder, and Barsha Gadapani Pathak

**Software:** Sarmila Mazumder, and Barsha Gadapani Pathak

**Supervision:** Sarmila Mazumder, and Gitau Mburu

**Validation:** Gitau Mburu and Ndema Habib

**Visualization:** Barsha Gadapani Pathak, Gitau Mburu and Ndema Habib

**Writing – original draft:** Barsha Gadapani Pathak.

**Writing – review & editing:** Barsha Gadapani Pathak, Gitau Mburu, Ndema Habib, Rita Kabra, James Kiarie, Ranadip Chowdhury, Neeta Dhabhai, and Sarmila Mazumder

## Data Availability Statement

The dataset used for this study is derived from a larger dataset collected for multiple research objectives. Only the data relevant to the specific research question addressed in this study has been extracted and analyzed. Due to ethical restrictions and confidentiality agreements, the full dataset is not publicly available. However, access to the data may be requested by contacting the Ethics Review Committee (ERC) at the Society for Applied Studies via email at or by phone at +91-7838350052. The ERC will review data access requests and provide guidance regarding any applicable restrictions.

## Acknowledgements

We thank the all the participants for their time to participate in this study. Additionally, authors thank the contribution of data manager Baljeet Kaur.

## Supporting information

**S1 Table:** Proportion of women seeking care for delayed pregnancy, source of care and treatment/management received.

**S 1 Fig:** Scree plot of eigen values after factor loading

**S 2 Table:** Multivariable model representing the determinants of care-seeking for delayed conception from all sources of care versus formal and mixed sources of care among study participants

